# Building and validating a predictive model for stroke risk in Chinese community-dwelling patients with chronic obstructive pulmonary disease using machine learning methods

**DOI:** 10.1101/2024.09.12.24313533

**Authors:** Yong Chen, Yonglin Yu, Dongmei Yang, Xiaoju Chen

## Abstract

**Background:** The occurrence of stroke in patients with chronic obstructive pulmonary disease (COPD) can have potentially devastating consequences; however, there is still a lack of predictive models that accurately predict the risk of stroke in community-based COPD patients in China. The aim of this study was to construct a novel predictive model that accurately predicts the predictive model for the risk of stroke in community-based COPD patients by applying a machine learning methodology within the Chinese community.

**Methods:** The clinical data of 809 Community COPD patients were analyzed by using the 2020 China Health and Retirement Longitudinal Study (CHARLS) database. The least absolute shrinkage and selection operator (LASSO) and multivariate logistic regression were used to analyze predictors. Multiple machine learning (ML) classification models are integrated to analyze and identify the optimal model, and Shapley Additive exPlanations (SHAP) interpretation was developed for personalized risk assessment.

**Results:** The following six variables:Heart_disease,Hyperlipidemia,Hypertension,ADL_score, Cesd_score and Parkinson are predictors of stroke in community-based COPD patients. Logistic classification model was the optimal model, test set area under curve (AUC) (95% confidence interval, CI):0.913 (0.835-0.992), accuracy: 0.823, sensitivity: 0.818, and specificity: 0.823.

**Conclusions:** The model constructed in this study has relatively reliable predictive performance, which helps clinical doctors identify high-risk populations of community COPD patients prone to stroke at an early stage.

## Introduction

Chronic obstructive pulmonary disease (COPD) is a common respiratory system disease that has a significant impact on the health status of patients worldwide. It is the third leading cause of death globally [1, 2]. The prevalence of COPD patients in the Chinese community remains high[3]. Among the complications associated with COPD, stroke is a serious condition that can decrease quality of life or lead to life-threatening situations[4]. Stroke, a disease caused by a sudden interruption of the blood supply to the brain or a rupture of a blood vessel in the brain. Strokes result in a lack of oxygen to the brain tissue, which impairs brain function and manifests itself in symptoms such as speech and motor deficits.COPD is more prone to strokes because it is often associated with systemic inflammation and oxidative stress[5, 6]. Therefore, timely prediction of the risk of stroke in community-based COPD patients is crucial for clinical intervention and treatment. Most of the past studies have been based on traditional statistical analysis methods to predict the risk of stroke in COPD patients, but these methods have certain limitations. These traditional statistical analysis methods usually assume that the relationship between variables is linear and require extensive manual feature extraction and selection, limiting the accuracy and efficiency of the prediction models. In recent years, the application of machine learning techniques has gradually attracted attention in the medical field. Machine learning is a method that utilizes computer algorithms to automatically learn and improve performance, which is able to handle large-scale and high-dimensional data and improve the accuracy and efficiency of prediction[7]. In addition, machine learning has the advantage of flexibility and scalability, and is able to solve the problem of multivariate interaction and covariance well. It builds risk models for predicting diseases, diagnosing disease severity, and evaluating disease prognosis by learning from data obtained from patients’ existing medical tests or surveys[8].In addition, there is currently no prediction model for the occurrence of stroke in COPD patients in the Chinese community. Therefore, the aim of this study is to use machine learning techniques to construct and validate a prediction model for stroke occurrence applicable to COPD patients in the Chinese community, and to compare the performance of different machine learning algorithms in this field. By training and testing a large amount of clinical data, we hope to develop a model that accurately predicts the risk of stroke in community-based COPD patients and provide a more effective risk prediction tool for clinical practice and better decision support for clinicians. This will help early intervention of stroke risk in community COPD patients and improve the quality of life and prognosis of patients.

## Materials and Methods

### Materials

The data utilized in this research were extracted from the 2020 China Health and Retirement Longitudinal Study (CHARLS) database, which can be accessed through the following link:(http://charls.pku.edu.cn). This research was granted approval by the Biomedical Ethics Committee at Peking University in Beijing (Approval No. IRB00001052-11015) and was conducted in accordance with the principles outlined in the Declaration of Helsinki. A total of 809 patients with chronic obstructive pulmonary disease were collected from the community.

### Inclusion and exclusion criteria

Inclusion criteria are as follows: (1)Patients diagnosed with COPD in the community who are aged ≥40; (2)Patients who have received at least one diagnosis of stroke during the study period; (3) Patients with complete clinical medical history records and relevant examination data.Exclusion criteria are as follows:(1)Patients with cognitive impairments or unable to provide reliable data;(2)Patients with other serious illnesses or complications that may affect the accuracy of stroke risk prediction; (3)Patients who fail to provide truthful information or data.

## Methods

The The study identified two distinct groups through a thorough assessment of the patients: those with COPD but no history of stroke, and those with COPD who had experienced stroke. The study examined data gathered from the 2020 CHARLS database, which included a total of 28 variables: Gender, Living condition,Education, Marital_status, Medical_insurance, Self_assessed_health , Hypertension, Diabetes, Hyperlipidemia, Tumours, Liver_disease, Heart_disease, Kidney_disease, Mental_disorder,Memory_disease,Parkinson, Heavy_physical_exercis, Mild_exercise, Moderate_exercise, Social_activities, go online , Smoking, Drinking, ADL_score , Cesd_score, lived_alone_days, living_with_partner_days and Age.Multiple interpolation filling was used for missing values to complete the data.Development and Assessment of Forecasting Models After selecting key factors from the independent variables, COPD patients were divided into a training set and testing set. Various ML classification models were utilized for in-depth analysis and comparison of the importance of each index in the training and testing sets among different models. The optimal model was then employed to validate and evaluate the results. Additionally, the SHAP presentation model, encompassing both an overall and single sample interpretation, was developed.

The detailed steps involved the following: (1) Screening key factors: Initially, R software was used to conduct a LASSO regression analysis to simplify variable selection and reduce complexity. Subsequently, the results of the LASSO regression analysis were utilized for multifactor logistic regression analysis using SPSS, leading to the identification of significant factors with a p-value of < 0.05. (2) Data partition: Python software utilized a random number method to randomly assign COPD patients into a training and test set in a 7:3 ratio.

Five machine learning algorithms, specifically logistic regression (LR), Support Vector Machine(SVM), eXtreme Gradient Boosting (XGBoost), random forest (RF), and Light Gradient Boosting Machine (LightGBM) were utilized to predict the risk of stroke in COPD patients. The training set employed k-fold cross-validation and a resampling approach with k=10. The validation set was used to optimize model parameters, and the test set was used to evaluate system performance. Model quality was evaluated using discrimination, calibration, and clinical utility measures, with calibration plots used to assess calibration and discrepancies between model predictions and actual events. Decision curve analysis (DCA) was utilized to determine the clinical benefit by calculating the net benefit of different probability thresholds. Confusion matrix metrics were used to evaluate mean precision, accuracy, sensitivity, specificity, and F-value scores of the models.It is important to recognize the limitations in interpreting results from machine learning techniques. The SHAP method, which is based on game theory, was implemented to interpret results from any machine learning model[9]. SHAP values were used to assess the importance of each predictor variable, with high values positively impacting the model output and low values having the opposite effect. Ultimately, a comprehensive analysis was conducted, incorporating seven variables.

### Statistical Analysis

In the analysis of the training and testing datasets, all variables were carefully considered. Continuous variables were described using their median and Interquartile Range (IQR) and analyzed using the Mann–Whitney U test. Categorical variables, on the other hand, were presented as the frequency and percentage of occurrences and compared using chi-square tests. Statistical significance was determined by two-tailed p-values below 0.05. The statistical analysis was conducted using SPSS (version 27.0), R (version 4.2.3), and Python (version 3.11.4).

## Results

A total of 809 community-based COPD patients were included in this study. Of these patients, 771 had no history of stroke, while 38 had a concurrent stroke(**table1**).

**Table 1.**
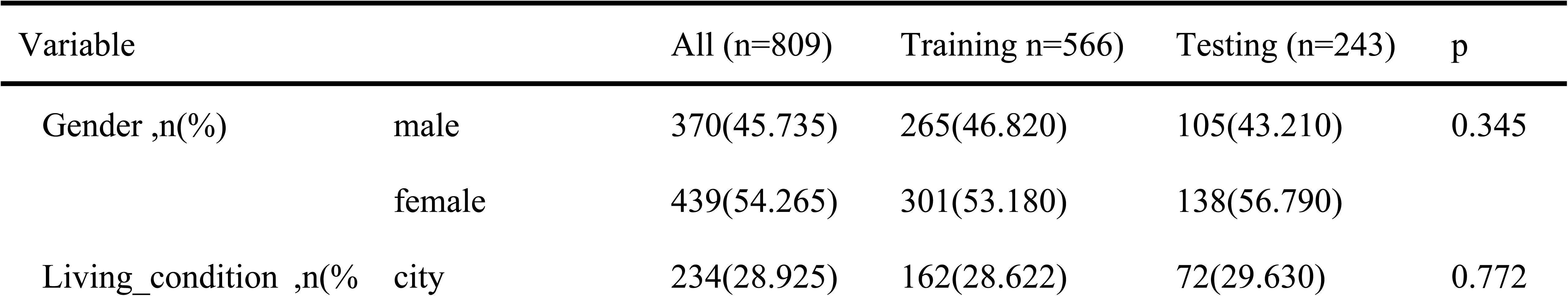
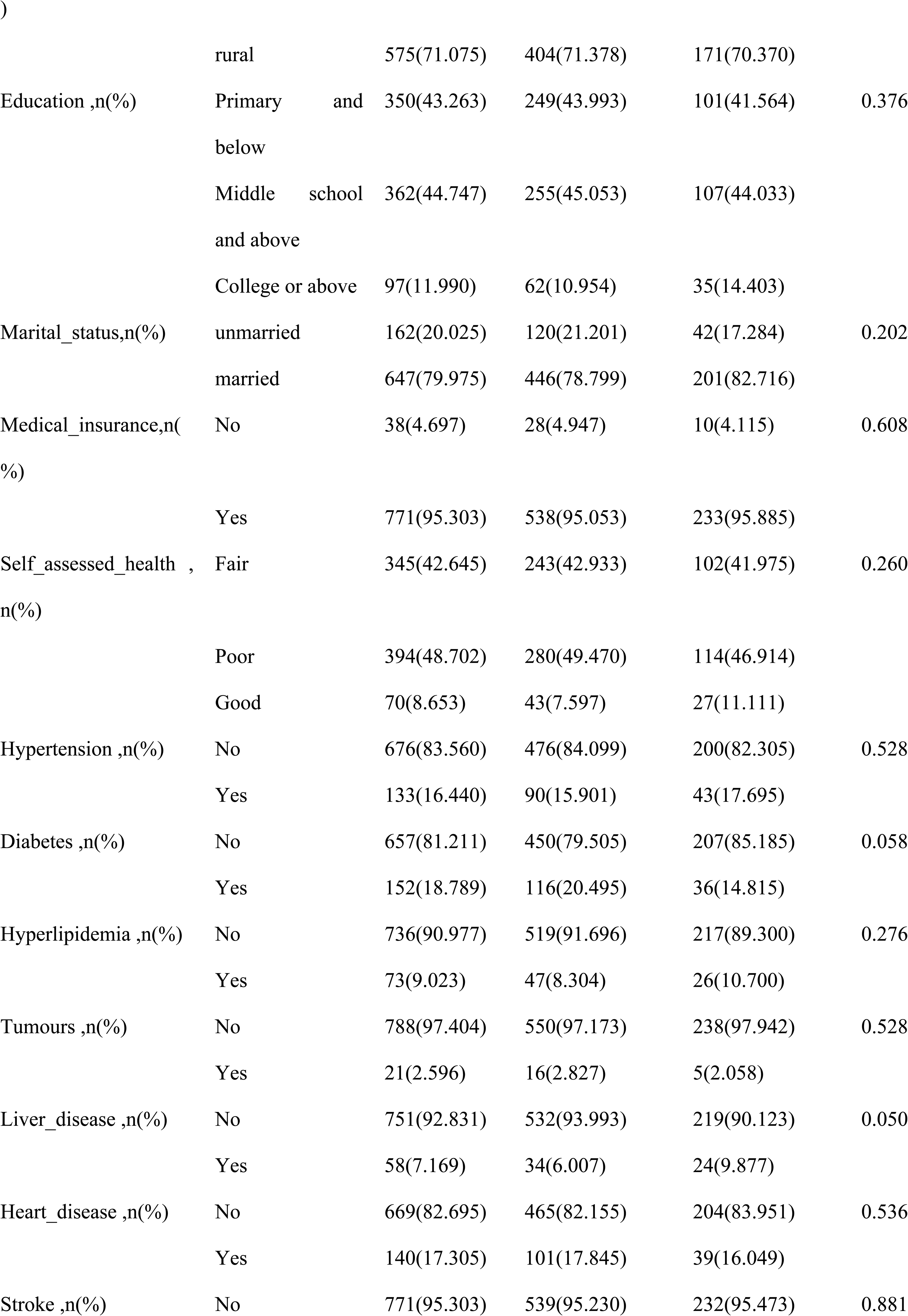
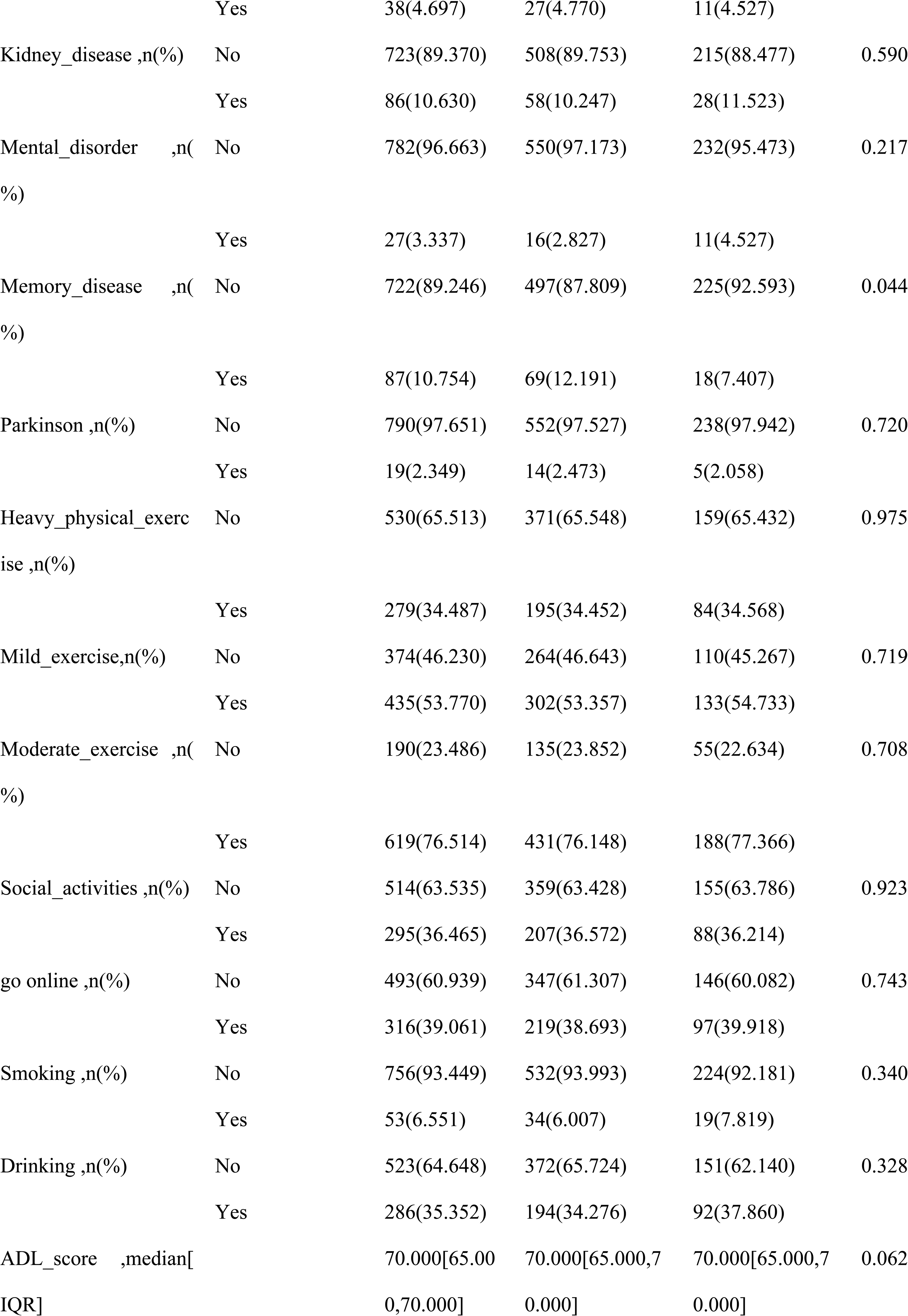
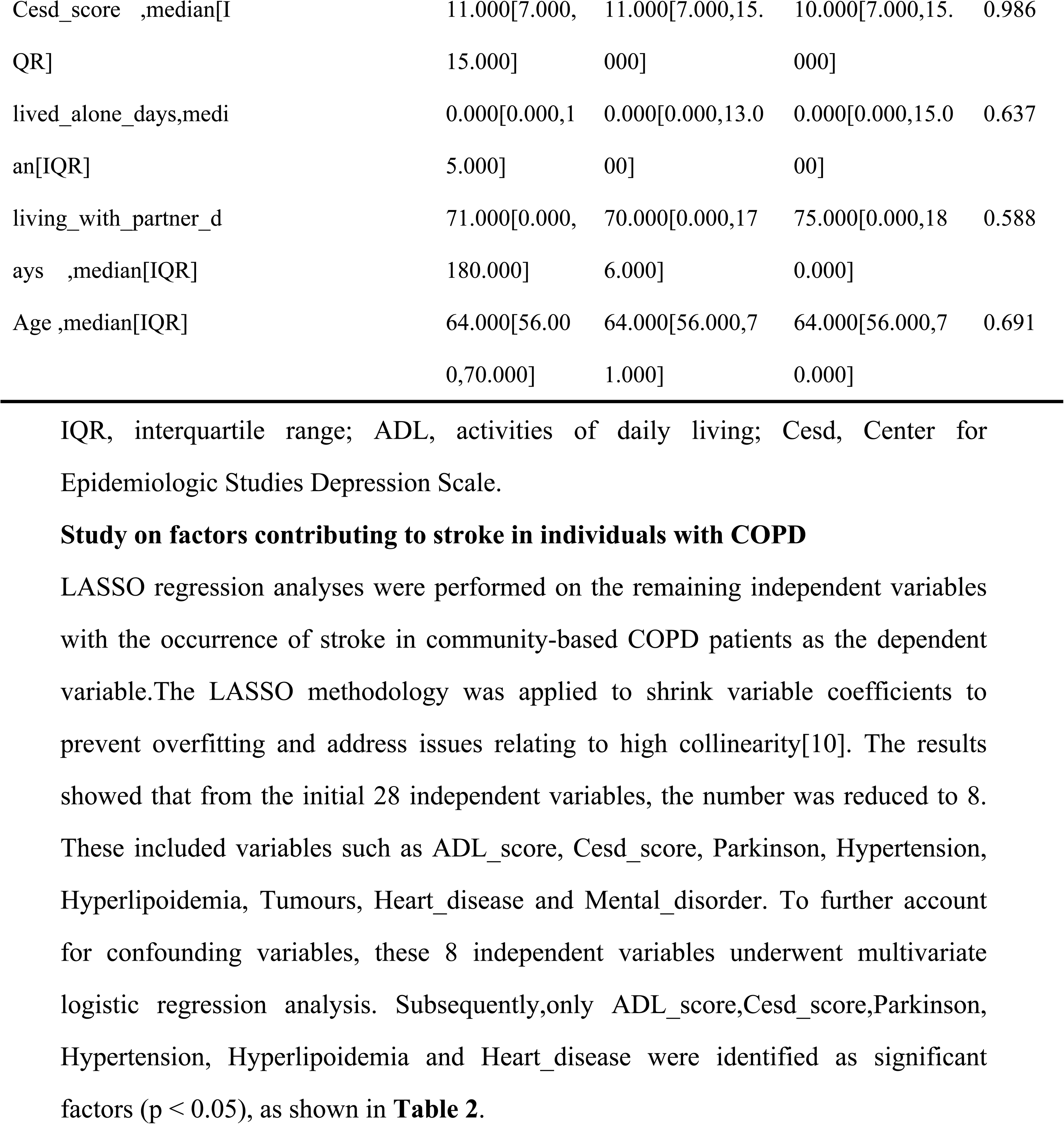
Baseline characteristics of the training cohort and testing cohort.

### Study on factors contributing to stroke in individuals with COPD

LASSO regression analyses were performed on the remaining independent variables with the occurrence of stroke in community-based COPD patients as the dependent variable.The LASSO methodology was applied to shrink variable coefficients to prevent overfitting and address issues relating to high collinearity[10]. The results showed that from the initial 28 independent variables, the number was reduced to 8. These included variables such as ADL_score, Cesd_score, Parkinson, Hypertension, Hyperlipoidemia, Tumours, Heart_disease and Mental_disorder. To further account for confounding variables, these 8 independent variables underwent multivariate logistic regression analysis. Subsequently,only ADL_score,Cesd_score,Parkinson, Hypertension, Hyperlipoidemia and Heart_disease were identified as significant factors (p < 0.05), as shown in **Table 2**.

**Table 2.** Multivariate logistic regression analysis.

| Variable | SE | Z | p | OR |
| --- | --- | --- | --- | --- |
| (Intercept) | 0.811 | -5.373 | 0.0 | 0.013(0.002-0.059) |
| ADL_score | 0.009 | -2.34 | 0.019 | 0.98(0.963-0.997) |
| Cesd_score | 0.035 | 2.575 | 0.01 | 1.094(1.022-1.173) |
| Parkinson | 0.715 | 1.998 | 0.046 | 4.173(0.965-16.326) |
| Hypertension | 0.468 | 2.371 | 0.018 | 3.03(1.189-7.535) |
| Hyperlipoidemia | 0.49 | 3.068 | 0.002 | 4.495(1.711-11.77) |
| Tumours | 0.843 | 1.785 | 0.074 | 4.503(0.688-20.132) |
| Heart_disease | 0.423 | 2.608 | 0.009 | 3.012(1.298-6.889) |
| Mental_disorder | 0.781 | 0.528 | 0.598 | 1.51(0.301-6.503) |

(SE, Standard error; OR, odds ratio; CI, confidence interval; )

### Thorough Examination of Categorized Multi-Model Analysis

Five ML methods, including LR, SVM, XGBoost, RF, and LightGBM were trained and iterated ten times. Area under the curve (AUC) values were used for the evaluation of the models[11]. XGBoost,RF and LightGBM had the highest AUC value in the training set (Fig 2A), while LR had the highest value in the validation set (Fig 2B). As AUC metrics focus on the predictive accuracy of the model and cannot indicate whether a model is clinically usable or determine which model may be preferable DCA, calibration curves, and precision-recall (PR) curves were used in this study[12]. DCA assessed the clinical applicability of the LR and XGBoost models in improving accuracy (Fig 2C). The XGBoost and LR model predictions were more accurate according to the calibration curves (Fig 2D). In the evaluation of the clinical applicability and prediction accuracy of the LR and RF models, the LR model showed the best performance in the training and validation sets, with the highest average precision (AP) values in the validation set (Fig 2E,F). A comprehensive analysis indicated that LR may be relatively stable considering the high probability of overfitting in RF, and thus LR was selected as the optimal model **(see more details in Supplemental Table S1)**.

**Fig 1.**
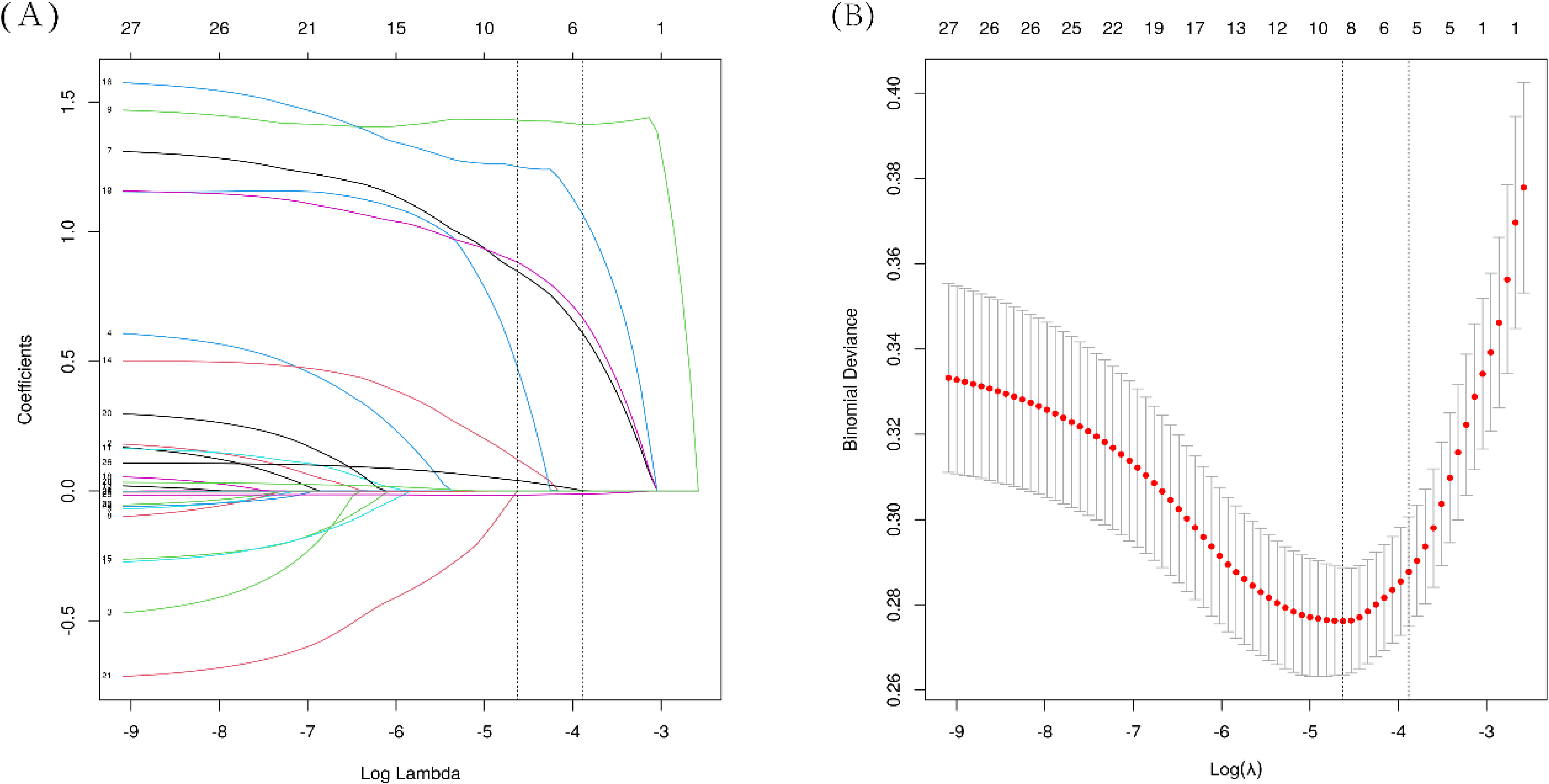
LASSO regression analysis was utilized to identify key factors. (A) Vertical lines delineating selected values were generated using 10-fold cross-validation, pinpointing the optimal lambda which yielded eight non-zero coefficients. (B) The LASSO model showcased coefficient profiles of 28 texture features plotted along the log(λ) sequence. Notably, vertical dotted lines were incorporated to highlight the minimum mean square error (λ = 0.01) and the standard error of the minimum distance (λ = 0.021).

**Fig 2.**
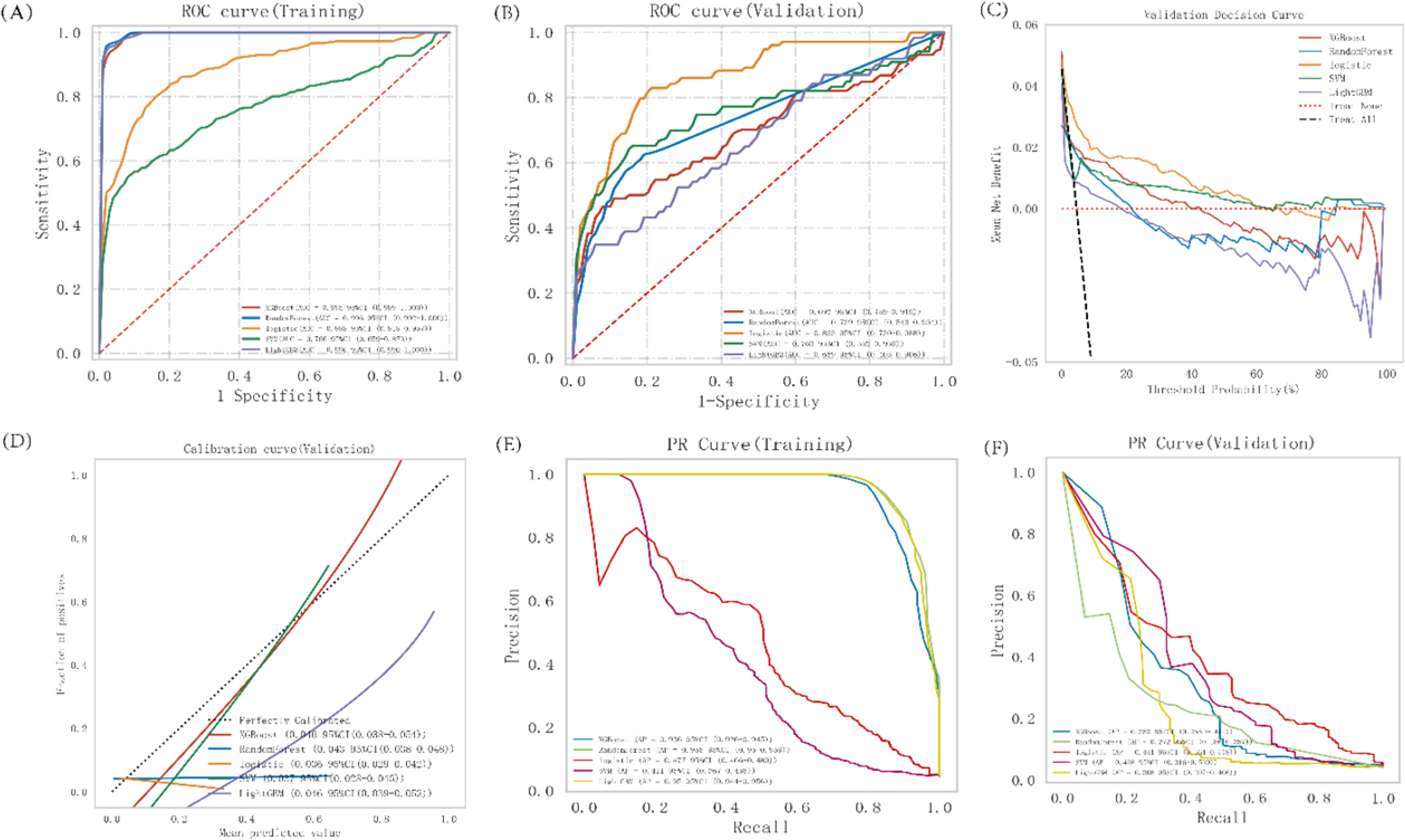
The ML model was evaluated through a comprehensive analysis. This included (A) ROC and AUC for the training set, and (B) ROC and AUC for the validation set.In the study, patients with COPD were sampled multiple times in a 7:3 ratio. (C) Validation set DCA showing the various model assumptions, with the black dashed line indicating that all patients had a stroke and the thin red and black lines indicating no stroke. Different models were represented by solid lines. (D) The calibration curve for the Verification set displayed the average prediction probability on the abscissa, actual probability of the event on the ordinate, and different model fitting lines compared to a reference line for accuracy assessment. (E) Included the PR curve and AP for the training set, while (F) illustrated the PR curve and AP for the verification set, with precision on the y-axis and recall on the x-axis. The PR curves of models were compared, with a model’s superiority indicated by one curve completely covering another. The higher the AP value, the better the model performance, with different colors representing each model and values displayed as averages with 95% CI.

### The Optimal Procedure for Constructing and Assessing Models

The dataset designated for training underwent LR analysis and 10-fold cross-validation. As a result, the training set yielded an average AUC (95% CI) of 0.865(0.774-0.957), while the average AUC from cross-validation of the validation set was 0.860(0.627-0.999). Moreover, the average AUC from the test set stood at 0.913 (0.835-0.992) (Fig 3A-C). The AUC values for the training set, validation set, and testing set were consistently stable at approximately 0.85. The model’s predictive performance was deemed to be highly accurate based on these results. The learning curves show strong consistency between the training and validation sets, indicating a high degree of fit and a high degree of stability[13, 14] (Fig 3D). These findings suggest that the Logistic regression model is suitable for classification modeling in this dataset. **(see more details in Supplemental Table S2)**.

**Fig 3.**
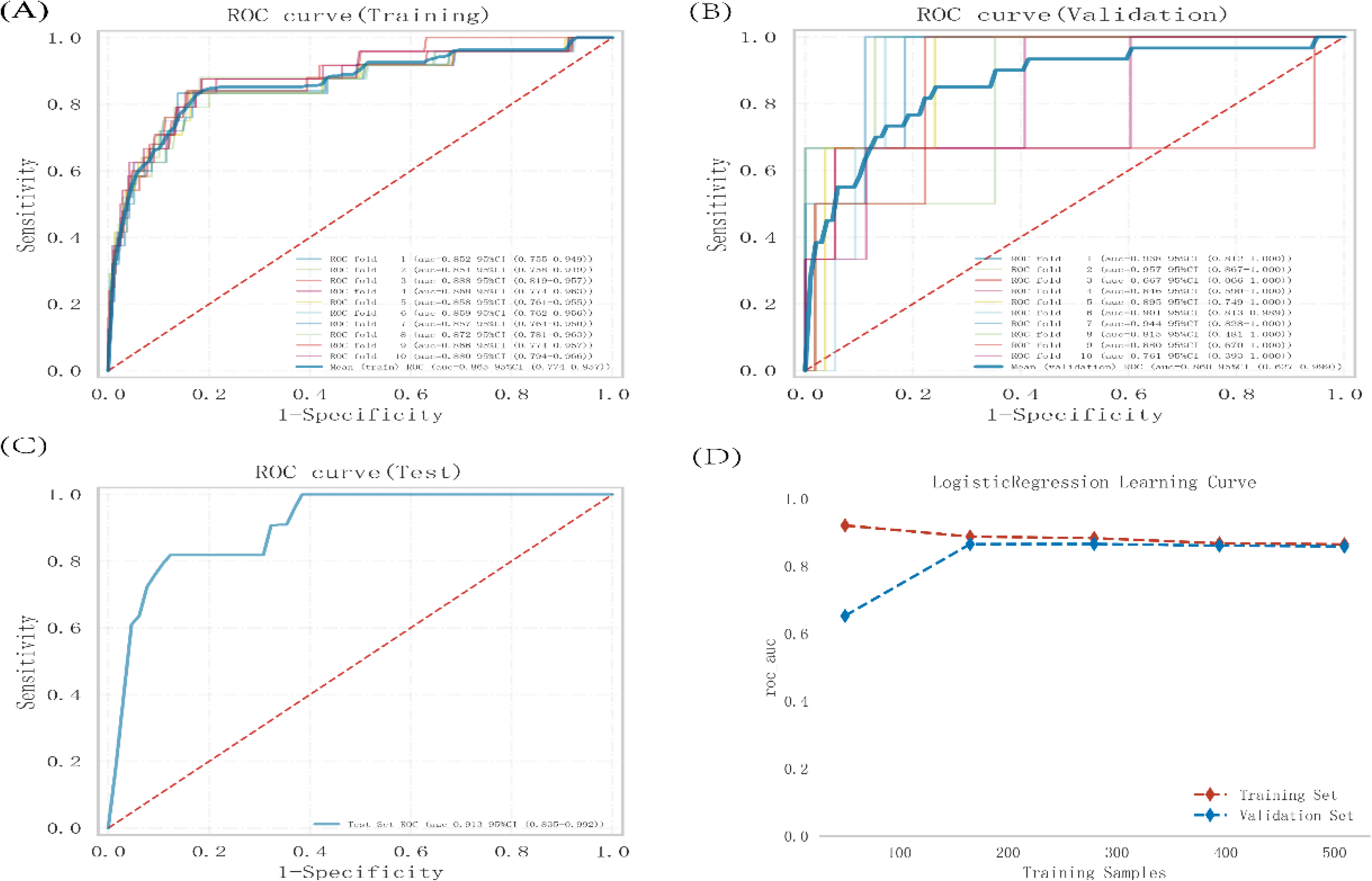
Logistic regression model training, validation, and testing. (A)Training sets ROC and AUC and (B)validation sets ROC and AUC. Training and cross-validation of 10% of COPD patients. Solid lines of different colors represent 10 different results. (C) Test set ROC and AUC. Test results for 30% of COPD patients. (D) Learning curve. The red dashed line represents the training set and the blue dashed line represents the validation set. The values are expressed in terms of average and 95% CI.

### The SHAP Approach to Interpreting Models

To visually illustrate the selected variables, we utilized SHAP to demonstrate how these variables influenced the prediction of a tophus in the model[9, 15]. (Fig 4A) displays the six most crucial features in our model. Each line representing an important feature showcases the attributions of all patients towards the outcomes, depicted by different colored dots – red dots signify high-risk values, while blue dots denote low-risk values. Factors such as: Heart_disease, Hyperlipidemia, Hypertension, ADL_score, Cesd_score, and Parkinson were identified as contributors to the occurrence of stroke in community-dwelling COPD patients. (Fig 4B) presents the ranking of six risk factors based on the average absolute SHAP value, with the x-axis SHAP value indicating the significance within the forecasting model. Furthermore, we have included a typical example in (Fig 4C) to emphasize the interpretability of the model.

**Fig 4.**
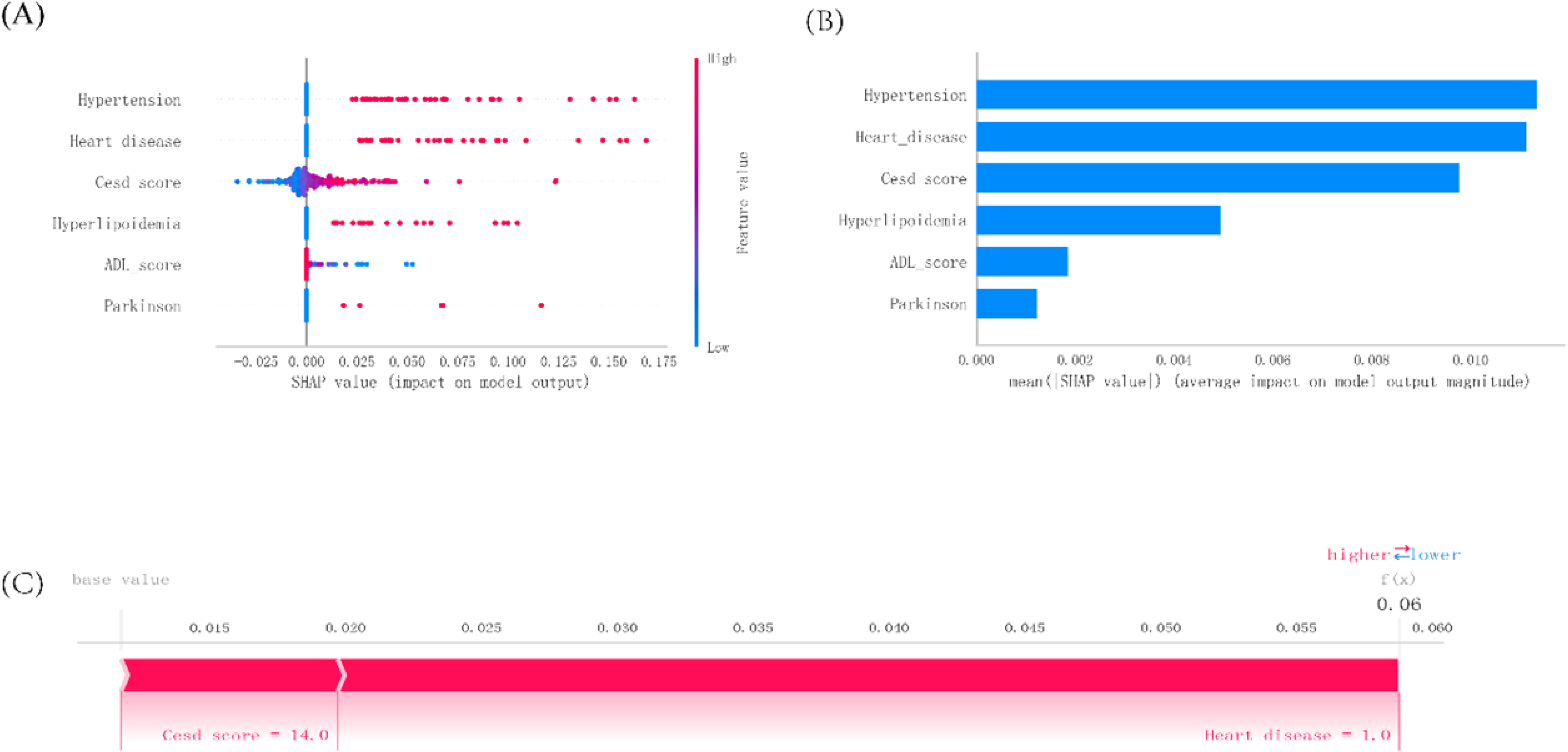
Explanation of the SHAP model. (A) Identifying key attributes through SHAP analysis. The x-axis represents the SHAP values, with each line corresponding to a specific feature. High values are denoted by red dots, while low values are denoted by blue dots. (B) Evaluation of feature importance based on SHAP analysis. This matrix diagram illustrates the contribution of each variable to the final predictive model. (C) Visualization of SHAP force plots. SHAP, also known as Shapley additive explanations, provides insights into the log odds ratio (F(x)) for each observation. The arrows indicate the impact of each factor on the prediction outcome, with blue arrows indicating a decrease in risk and red arrows indicating an increase in risk of stroke. The length of the arrow correlates with the magnitude of the effect. The length of the arrow correlates with the magnitude of the effect.

## Discussion

Chronic Obstructive Pulmonary Disease (COPD) remains a significant global health burden with increasing prevalence, especially in aging populations and in areas with high prevalence of smoking and air pollution [16]. Stroke is the leading cause of death and disability worldwide, and not only may result in long-term disability, but can also incur high associated treatment costs[17, 18]. In addition, it has been found that the prevalence of both COPD and stroke is high, and both are common chronic diseases, which not only seriously affect the quality of life of patients, but also impose a great economic burden on society and families[19–21]. In this study, we retrospectively analyzed the indicators of Gender, Living condition, Education, Marital_status, Medical_insurance, Self_assessed_health , Hypertension, Diabetes, Hyperlipidemia, Tumours, Liver_disease, Heart_disease, Kidney_disease, Mental_disorder, Memory_disease, Parkinson, Heavy_physical_exercis, Mild_exercise, Moderate_exercise, Social_activities, go online, Smoking, Drinking, ADL_score, Cesd_score, lived_alone_days, living_with_partner_days, and Age in community COPD patients who had stroke, and after analyzing by LASSO and multivariate logistic regression, we concluded that the combination of Heart_disease, Hyperlipidemia, Hypertension, ADL_score, Cesd_score, and Parkinson are risk factors for stroke in community-based COPD patients.

COPD and heart disease frequently co-occur and are associated with worse prognostic outcomes [22]. In turn, heart disease and stroke attack share common risk factors, and previous studies have found [23] that cardiovascular disease and cerebrovascular disease share common pathogenesis and risk factors. Therefore, some studies have found that COPD combined with related heart diseases such as [24, 25] : atrial fibrillation, myocardial infarction, coronary artery disease, heart failure, and hypertension are more prone to stroke, and they hypothesized that it may be coexisting with having similar risk factors such as aging, smoking history, sedentary lifestyle, and systemic inflammatory response [26]. Our study showed a significant correlation between hypertension and stroke outcome in community-based COPD patients. This is also in line with existing studies [4, 27]. The possible mechanisms are [26, 28]: the higher pressure on the vessel walls in hypertensive patients leads to thinning of the vessel walls in the long term, and the fragile vessels are prone to rupture, resulting in hemorrhagic stroke. In addition, hypertension accelerates the process of atherosclerosis, narrowing or occluding the cerebral blood vessels, thus increasing the risk of ischemic stroke.COPD patients may also suffer from endothelial damage due to prolonged hypoxia and inflammatory response, further aggravating vascular pathology.Increased inflammatory response in COPD patients may result in unstable atherosclerotic plaques, which are prone to rupture and form thrombi, blocking the cerebral blood vessels This can lead to stroke. At the same time, inflammatory factors also affect blood pressure control, making the condition of hypertensive patients more serious. Therefore, there is a close link between stroke and hypertension in patients with COPD, and they interact with each other through common risk factors, hypoxia, inflammatory response, vascular remodeling, and other mechanisms.

The results of the present study suggest that patients with COPD in the community have an elevated risk of stroke in combination with hyperlipidemia.COPD and hyperlipidemia tend to co-exist, which may be associated with similar risk factors, such as smoking, lack of exercise, and poor dietary habits[29]. At the same time, the coexistence of these two diseases exacerbates systemic inflammation and metabolic disturbances, multiplying the risk of stroke[28, 30]. Patients with COPD typically experience oxidative stress, with increased production of reactive oxygen species (ROS) due to inflammation, which can damage endothelial cells and promote lipid peroxidation[31]. This damage can accelerate the formation of atherosclerosis and may also affect lipid metabolism, further contributing to the exacerbation of hyperlipidemia.Patients with COPD are often accompanied by hypoxemia, and prolonged hypoxia also activates certain metabolic pathways, leading to abnormalities in lipid metabolism, exacerbating hyperlipidemia, and further increasing the risk of stroke[32]. It has been found that certain medications used to treat COPD may affect lipid metabolism and increase blood lipid levels [4]. For example, long-term steroid use may lead to weight gain and lipid metabolism disorders, further promoting the development of hyperlipidemia.

In addition, the present study’s also found that community-based COPD patients with comorbid Parkinson’s disease were also prone to stroke.COPD and Parkinson’s disease are commonly found in middle-aged and elderly populations, and the prevalence of both diseases increases with age[33]. The aging process includes a variety of pathological changes, such as abnormal cell signaling pathways and decreased antioxidant capacity, which may affect the development of both COPD and Parkinson’s disease. Patients with Parkinson’s disease usually suffer from dyskinesia, resulting in decreased physical activity[34]. It has been found that lack of exercise not only exacerbates the symptoms of COPD, but may also lead to deterioration of cardiovascular health, which may increase the risk of stroke[35]. On the other hand, COPD patients with limited ability to perform daily activities due to dyspnea may likewise develop dyskinesia, increasing the overall risk of stroke. It has also been suggested that both COPD and Parkinson’s disease are associated with chronic inflammation[36]. Chronic inflammation in COPD patients leads to elevated levels of systemic inflammation, which may have an impact on the nervous system through pro-inflammatory cytokines, thereby increasing the risk of neurodegenerative diseases. In addition, inflammation may affect the integrity of the blood-brain barrier, making it easier for inflammatory factors to enter the central nervous system and exacerbate the course of Parkinson’s disease . In addition oxidative stress in chronic obstructive airway disease has been associated with Parkinson’s disease[37].The above through mechanisms such as inflammatory response thereby triggering the combination of Parkinson’s disease in patients with chronic obstructive pulmonary disease are more prone to stroke. A study on ACOS [38] found a higher risk of stroke and Parkinson’s in patients with ACOS. There are fewer studies directly on the occurrence of stroke in COPD combined with Parkinson’s disease and more studies are needed.

Our study further found a significant correlation between the occurrence of stroke and activities of daily living (ADL) scores in community-based COPD patients. This correlation can be explored through a variety of mechanisms, as follows: patients with COPD often suffer from decreased mobility due to dyspnea and limited physical activity, which affects their ADL[39, 40]. Low ADL_scores are usually associated with high stroke risk. It is because low mobility may lead to muscle atrophy, deterioration of cardiovascular health and metabolic disorders, which in turn increase the incidence of stroke. It has been found that chronic inflammation accompanying COPD can trigger a systemic response[41]. There is an association between chronic inflammation and decreased ADL_scores[42, 43], and lower ADL_scores may reflect an increased systemic inflammatory burden, which can be associated with stroke risk. In addition, COPD patients are often in a state of oxidative stress, which can cause damage to multiple organ systems, including the cardiovascular system. The oxidative stress may also lead to muscle fatigue and functional decline, further reducing ADL_ scores[44]. The occurrence of stroke is associated with oxidative stress, and therefore, a reduced ADL_score may indirectly reflect the risk of stroke.Patients with COPD may face mental health problems, such as depression and anxiety, and these psychological states can negatively affect daily activities, which can reduce ADL_ scores[45]. In turn, low ADL_scores can affect patients’ self-perception and social engagement, increasing the psychological and physical burden of stroke risk.

It also follows from within our research that the occurrence of stroke in community-based COPD patients is associated with Cesd_score. It was found that the Cesd_score is a tool used to assess depressive symptoms and that higher Cesd_score usually imply increased severity of depressive symptoms[46]. The depressive symptoms affect an individual’s physiological state, including immune function, inflammatory response, and cardiovascular health[47]. An elevated Cesd_score is usually associated with lifestyle changes and decreased self-management. The depressive mood may lead to conditions such as reduced physical activity, poor diet and failure to take medications on time. This lifestyle change not only affects symptom control in COPD patients, but may also lead to deterioration of cardiovascular health, which may increase the risk of stroke[35]. In addition, depressive symptoms often lead to decreased participation in social activities, resulting in increased loneliness.COPD patients who reduce their social activities due to depression may have an impact on their cardiovascular health and blood supply to the brain, which may increase the risk of stroke[48–50]. In conclusion, the relationship between the occurrence of stroke and Cesd_score in COPD patients is complex and involves psychological, physiological, and social factors. Therefore, attention should be paid to the mental health of COPD patients in clinical management, with early assessment and intervention for depressive symptoms to reduce the risk of stroke and improve overall quality of life.

### Limitation

However, it is important to recognize the limitations of our study. First, the lack of universally accepted criteria for inclusion or exclusion of a factor is an obvious constraint. Second, the sample size used in the study was relatively small, thus limiting the applicability of the results. Although the results of the training and test sets were analyzed to a high level of agreement, there is still the potential for error due to uncertainty in the criteria chosen. In addition, certain variables such as alcohol consumption and diabetes were not considered in the study design. Clearly, further longitudinal or prospective case-control studies are necessary to elucidate the correlation between risk factors and stroke incidence in community-based COPD patients.

### Conclusions

In conclusion, we constructed a predictive model based on the ML model, and the logistic regression model showed better performance in this study. In addition, we provided a personalized risk assessment for preventing the occurrence of stroke in COPD patients in the community, which was interpreted by SHAP. This effective computer-assisted method can help frontline clinicians and patients to recognize and intervene in the occurrence of stroke.

## Data Availability

The datasets utilized in this study can be obtained from the corresponding author upon submission of a reasonable request.

## Abbreviations

COPD: chronic obstructive pulmonary disease
LASSO: Least Absolute Shrinkage and Selection Operator
AUC: area under the receiver operating characteristics curve
DCA: decision curve analysis
ADL: activity of daily living
OR: Odds Ratio
*CI*: Confidence Interval

## Ethics Approval and Informed Consent Ethical

The research that included human participants underwent review and approval by CHARLS, as it was ethically approved by the Ethics Review Board of Peking University with (approval number IRB00001052-11015). Each participant provided their consent by signing an informed consent form. This study did not require written informed consent for participation as per the national laws and institutional regulations.

## Consent for Publication

All authors have given their consent for the publication of this work.

## Author Contributions

As co-first authors, Yong Chen and Yonglin Yu have equally contributed to this study. Yong Chen and Dongmei Yang conceptualized and designed the study. Formal analysis, initial drafting, and the idea were carried out by Yonglin Yu and Xiaoju Chen. Investigation oversight was provided by Yong Chen and Yonglin Yu. The paper was authored by Yong Chen and Dongmei Yang, while resource management and data curation were overseen by Dongmei Yang.

## References

1. Kahnert K, Jörres RA, Behr J, Welte T: The Diagnosis and Treatment of COPD and Its Comorbidities. Dtsch Arztebl Int 2023, 120(25):434–444.

2. Safiri S, Carson-Chahhoud K, Noori M, Nejadghaderi SA, Sullman MJM, Ahmadian Heris J, Ansarin K, Mansournia MA, Collins GS, Kolahi AA et al: Burden of chronic obstructive pulmonary disease and its attributable risk factors in 204 countries and territories, 1990-2019: results from the Global Burden of Disease Study 2019. Bmj 2022, 378:e069679.

3. Wang C, Xu J, Yang L, Xu Y, Zhang X, Bai C, Kang J, Ran P, Shen H, Wen F et al: Prevalence and risk factors of chronic obstructive pulmonary disease in China (the China Pulmonary Health [CPH] study): a national cross-sectional study. Lancet 2018, 391(10131):1706–1717.

4. Shen AL, Lin HL, Lin HC, Chao JC, Hsu CY, Chen CY: The effects of medications for treating COPD and allied conditions on stroke: a population-based cohort study. NPJ Prim Care Respir Med 2022, 32(1):4.

5. Lahousse L, Tiemeier H, Ikram MA, Brusselle GG: Chronic obstructive pulmonary disease and cerebrovascular disease: A comprehensive review. Respir Med 2015, 109(11):1371–1380.

6. Ding C, Wang R, Gong X, Yuan Y: Stroke risk of COPD patients and death risk of COPD patients following a stroke: A systematic review and meta-analysis. Medicine (Baltimore) 2023, 102(47):e35502.

7. Choi RY, Coyner AS, Kalpathy-Cramer J, Chiang MF, Campbell JP: Introduction to Machine Learning, Neural Networks, and Deep Learning. Transl Vis Sci Technol 2020, 9(2):14.

8. Murdoch TB, Detsky AS: The inevitable application of big data to health care. Jama 2013, 309(13):1351–1352.

9. Lundberg SM, Erion G, Chen H, DeGrave A, Prutkin JM, Nair B, Katz R, Himmelfarb J, Bansal N, Lee SI: From Local Explanations to Global Understanding with Explainable AI for Trees. Nat Mach Intell 2020, 2(1):56–67.

10. Sauerbrei W, Royston P, Binder H: Selection of important variables and determination of functional form for continuous predictors in multivariable model building. Stat Med 2007, 26(30):5512–5528.

11. Obuchowski NA, Bullen JA: Receiver operating characteristic (ROC) curves: review of methods with applications in diagnostic medicine. Phys Med Biol 2018, 63(7):07tr01.

12. Dankers F, Traverso A, Wee L, van Kuijk SMJ: Prediction Modeling Methodology. In: Fundamentals of Clinical Data Science. edn. Edited by Kubben P, Dumontier M, Dekker A. Cham (CH): Springer Copyright 2019, The Author(s). 2019: 101–120.

13. Belkin M, Hsu D, Ma S, Mandal S: Reconciling modern machine-learning practice and the classical bias-variance trade-off. Proc Natl Acad Sci U S A 2019, 116(32):15849–15854.

14. Kernbach JM, Staartjes VE: Foundations of Machine Learning-Based Clinical Prediction Modeling: Part I-Introduction and General Principles. Acta Neurochir Suppl 2022, 134:7–13.

15. Gramegna A, Giudici P: SHAP and LIME: An Evaluation of Discriminative Power in Credit Risk. Front Artif Intell 2021, 4:752558.

16. Li CL, Liu SF: Exploring Molecular Mechanisms and Biomarkers in COPD: An Overview of Current Advancements and Perspectives. Int J Mol Sci 2024, 25(13).

17. Tan KS, Pandian JD, Liu L, Toyoda K, Leung TWH, Uchiyama S, Kuroda S, Suwanwela NC, Aaron S, Chang HM et al: Stroke in Asia. Cerebrovasc Dis Extra 2024, 14(1):58–75.

18. Roth GA, Mensah GA, Johnson CO, Addolorato G, Ammirati E, Baddour LM, Barengo NC, Beaton AZ, Benjamin EJ, Benziger CP et al: Global Burden of Cardiovascular Diseases and Risk Factors, 1990-2019: Update From the GBD 2019 Study. J Am Coll Cardiol 2020, 76(25):2982–3021.

19. Zhou M, Wang H, Zeng X, Yin P, Zhu J, Chen W, Li X, Wang L, Wang L, Liu Y et al: Mortality, morbidity, and risk factors in China and its provinces, 1990-2017: a systematic analysis for the Global Burden of Disease Study 2017. Lancet 2019, 394(10204):1145–1158.

20. Burden of disease scenarios for 204 countries and territories, 2022-2050: a forecasting analysis for the Global Burden of Disease Study 2021. Lancet 2024, 403(10440):2204–2256.

21. Lackland DT, Roccella EJ, Deutsch AF, Fornage M, George MG, Howard G, Kissela BM, Kittner SJ, Lichtman JH, Lisabeth LD et al: Factors influencing the decline in stroke mortality: a statement from the American Heart Association/American Stroke Association. Stroke 2014, 45(1):315–353.

22. “Cardiovascular disease and COPD: dangerous liaisons.“ Klaus F. Rabe, John R. Hurst and Samy Suissa. Eur Respir Rev 2018; 27: 180057. *Eur Respir Rev* 2018, 27(150).

23. Han CH, Kim H, Lee S, Chung JH: Knowledge and Poor Understanding Factors of Stroke and Heart Attack Symptoms. Int J Environ Res Public Health 2019, 16(19).

24. Putcha N, Puhan MA, Hansel NN, Drummond MB, Boyd CM: Impact of co-morbidities on self-rated health in self-reported COPD: an analysis of NHANES 2001-2008. Copd 2013, 10(3):324–332.

25. Divo M, Cote C, de Torres JP, Casanova C, Marin JM, Pinto-Plata V, Zulueta J, Cabrera C, Zagaceta J, Hunninghake G et al: Comorbidities and risk of mortality in patients with chronic obstructive pulmonary disease. Am J Respir Crit Care Med 2012, 186(2):155–161.

26. Corlateanu A, Covantev S, Mathioudakis AG, Botnaru V, Cazzola M, Siafakas N: Chronic Obstructive Pulmonary Disease and Stroke. Copd 2018, 15(4):405–413.

27. Chen YF, Cheng YC, Chou CH, Chen CY, Yu CJ: Major comorbidities lead to the risk of adverse cardiovascular events in chronic obstructive pulmonary disease patients using inhaled long-acting bronchodilators: a case-control study. BMC Pulm Med 2019, 19(1):233.

28. Austin V, Crack PJ, Bozinovski S, Miller AA, Vlahos R: COPD and stroke: are systemic inflammation and oxidative stress the missing links? Clin Sci (Lond) 2016, 130(13):1039–1050.

29. Barr RG, Celli BR, Mannino DM, Petty T, Rennard SI, Sciurba FC, Stoller JK, Thomashow BM, Turino GM: Comorbidities, patient knowledge, and disease management in a national sample of patients with COPD. Am J Med 2009, 122(4):348–355.

30. Corlateanu A, Covantev S, Mathioudakis AG, Botnaru V, Siafakas N: Prevalence and burden of comorbidities in Chronic Obstructive Pulmonary Disease. Respir Investig 2016, 54(6):387–396.

31. Kaźmierczak M, Ciebiada M, Pękala-Wojciechowska A, Pawłowski M, Nielepkowicz-Goździńska A, Antczak A: Evaluation of Markers of Inflammation and Oxidative Stress in COPD Patients with or without Cardiovascular Comorbidities. Heart Lung Circ 2015, 24(8):817–823.

32. Geltser BI, Kurpatov IG, Kotelnikov VN, Zayats YV: Chronic obstructive pulmonary disease and cerebrovascular diseases: functional and clinical aspect of comorbidity. Ter Arkh 2018, 90(3):81–88.

33. Li CH, Chen WC, Liao WC, Tu CY, Lin CL, Sung FC, Chen CH, Hsu WH: The association between chronic obstructive pulmonary disease and Parkinson’s disease: a nationwide population-based retrospective cohort study. Qjm 2015, 108(1):39–45.

34. Gerlach OH, Broen MP, van Domburg PH, Vermeij AJ, Weber WE: Deterioration of Parkinson’s disease during hospitalization: survey of 684 patients. BMC Neurol 2012, 12:13.

35. Dugravot A, Fayosse A, Dumurgier J, Bouillon K, Rayana TB, Schnitzler A, Kivimaki M, Sabia S, Singh-Manoux A: Social inequalities in multimorbidity, frailty, disability, and transitions to mortality: a 24-year follow-up of the Whitehall II cohort study. Lancet Public Health 2020, 5(1):e42–e50.

36. Huang YF, Yeh CC, Chou YC, Hu CJ, Cherng YG, Shih CC, Chen TL, Liao CC: Stroke in Parkinson’s disease. Qjm 2019, 112(4):269–274.

37. Rusanen M, Ngandu T, Laatikainen T, Tuomilehto J, Soininen H, Kivipelto M: Chronic obstructive pulmonary disease and asthma and the risk of mild cognitive impairment and dementia: a population based CAIDE study. Curr Alzheimer Res 2013, 10(5):549–555.

38. Yeh JJ, Wei YF, Lin CL, Hsu WH: Effect of the asthma-chronic obstructive pulmonary disease syndrome on the stroke, Parkinson’s disease, and dementia: a national cohort study. Oncotarget 2018, 9(15):12418–12431.

39. Ozsoy I, Ozcan Kahraman B, Acar S, Ozalevli S, Akkoclu A, Savci S: Factors Influencing Activities of Daily Living in Subjects With COPD. Respir Care 2019, 64(2):189–195.

40. Lahaije AJ, van Helvoort HA, Dekhuijzen PN, Heijdra YF: Physiologic limitations during daily life activities in COPD patients. Respir Med 2010, 104(8):1152–1159.

41. Xu J, Zeng Q, Li S, Su Q, Fan H: Inflammation mechanism and research progress of COPD. Front Immunol 2024, 15:1404615.

42. Qin K, Lin L, Lu C, Chen W, Guo VY: Association between systemic inflammation and activities of daily living disability among Chinese elderly individuals: the mediating role of handgrip strength. Aging Clin Exp Res 2022, 34(4):767–774.

43. Lima-Costa MF, Mambrini JVM, Torres KCL, Peixoto SV, Andrade FB, De Oliveira C, Tarazona-Santos E, Teixeira-Carvalho A, Martins-Filho OA: Multiple inflammatory markers and 15-year incident ADL disability in admixed older adults: The Bambui-Epigen Study. Arch Gerontol Geriatr 2017, 72:103–107.

44. Li X, Cao X, Ying Z, Yang G, Hoogendijk EO, Liu Z: Plasma superoxide dismutase activity in relation to disability in activities of daily living and objective physical functioning among Chinese older adults. Maturitas 2022, 161:12–17.

45. Kanervisto M, Saarelainen S, Vasankari T, Jousilahti P, Heistaro S, Heliövaara M, Luukkaala T, Paavilainen E: COPD, chronic bronchitis and capacity for day-to-day activities: negative impact of illness on the health-related quality of life. Chron Respir Dis 2010, 7(4):207–215.

46. Zhou L, Ma X, Wang W: Relationship between Cognitive Performance and Depressive Symptoms in Chinese Older Adults: The China Health and Retirement Longitudinal Study (CHARLS). J Affect Disord 2021, 281:454–458.

47. Beurel E, Toups M, Nemeroff CB: The Bidirectional Relationship of Depression and Inflammation: Double Trouble. Neuron 2020, 107(2):234–256.

48. Sigurgeirsdottir J, Halldorsdottir S, Arnardottir RH, Gudmundsson G, Bjornsson EH: COPD patients’ experiences, self-reported needs, and needs-driven strategies to cope with self-management. Int J Chron Obstruct Pulmon Dis 2019, 14:1033–1043.

49. O’Donnell DE, Milne KM, James MD, de Torres JP, Neder JA: Dyspnea in COPD: New Mechanistic Insights and Management Implications. Adv Ther 2020, 37(1):41–60.

50. Portegies ML, Lahousse L, Joos GF, Hofman A, Koudstaal PJ, Stricker BH, Brusselle GG, Ikram MA: Chronic Obstructive Pulmonary Disease and the Risk of Stroke. The Rotterdam Study. Am J Respir Crit Care Med 2016, 193(3):251–258.

